# MRI-based volume assessments show no changes in hippocampus, amygdala, thalamus and brainstem subregions in narcolepsy type 1

**DOI:** 10.64898/2026.02.21.26345265

**Authors:** Hilde T. Juvodden, Dag Alnæs, Ingrid Agartz, Ole A. Andreassen, Andres Server, Per M. Thorsby, Lars T. Westlye, Stine Knudsen-Heier

## Abstract

**Study Objectives:** Narcolepsy type 1 (NT1) is characterized by excessive daytime sleepiness and cataplexy. Previous studies have implicated the amygdala, thalamus, brainstem and hippocampus in the pathophysiology of NT1. We here aimed to examine more detailed subregional case-control differences in MRI-based segmentations of these brain regions to gain deeper insights.

**Methods:** We obtained 3T MRI brain scans from 54 NT1 patients (39 females, mean age 21.8 ± 11.0 years, 51 with confirmed hypocretin-deficiency and three patients that had not performed this measure) and 114 healthy controls (77 females, mean age 23.2 ± 9.0 years). Automated segmentation of the hippocampus, amygdala, thalamus, and brainstem was performed on T1-weighted MRI data using FreeSurfer. Case-control volume differences were tested using general linear models and permutation testing. The false discovery rate was controlled at 5% with the Benjamini-Hochberg procedure.

**Results:** The analysis revealed no significant case-control differences for any of the subregions in the hippocampus, thalamus, amygdala and brainstem after correction for multiple testing.

**Conclusions:** Based on a detailed automated MRI-based segmentation analyses in a relatively large national sample, NT1 patients had no significant changes in any amygdala, thalamus, brainstem or hippocampus subregions compared to controls. In the future large multi-site studies could be performed to achieve sufficient power to detect more subtle group differences.

## 1. Introduction

The neurological sleep disorder narcolepsy type 1 (NT1) is characterized by excessive daytime sleepiness and cataplexy [1]. Cataplexy is considered the most pathognomonic symptom of NT1 and is characterized by sudden loss of muscle tone often elicited by strong positive emotions [2]. NT1 patients often also have disturbed nocturnal sleep and can experience hypnagogic (occur at sleep onset) or hypnopompic (on awakening) hallucinations and sleep paralysis [1]. After the mass vaccination in Norway and several other countries in 2009/2010 with Pandemrix®, there was over a 10-fold increase in new cases of NT1 [3, 4].

NT1 patients have loss/dysfunction of hypothalamic hypocretin-producing neurons, which widely projects to other brain regions, [1, 5, 6] and play an important role in the normal brain regulation/stabilization of sleep/wakefulness [7]. The hypothalamus has been extensively studied in NT1 and the loss/dysfunction of the hypocretin-producing neurons is most likely due to an auto-immune process. [1, 5] However, other brain regions have also been implicated in NT1. This includes the amygdala, which is considered part of the neurocircuitry involved in cataplexy [2, 8], the thalamus and the brainstem, which are important in the sleep-wake circuitry, including several nuclei in the pons, which constitute parts of the arousal system [7]. Despite conflicting evidence of memory impairments in NT1 [9], deficits in memory consolidation of motor and visual discrimination skills have been reported in NT1 patients [10].

Most previous MRI studies investigating NT1 case-control differences in subcortical volumes (including thalamus, hippocampus, amygdala or the brainstem) [11–19] have considered the total volume and not the corresponding more detailed subregional segmentations to explore if there are subregional volume differences within thalamus, hippocampus, amygdala or the brainstem. One study reported smaller total amygdala volume in NT1 patients compared to controls [11], while other reported no case-control differences [12–15]. Some MRI studies have reported smaller total hippocampus volume in NT1 patients compared to controls [15, 20], while others reported no difference [12, 14] or larger total hippocampus volume in NT1 patients [21]. Further, while smaller total thalamus volume in NT1 patients have been reported [12, 14], other studies reported no significant case-control differences in the amygdala, hippocampus and thalamus [16–19].

Manual segmentations can be time-consuming and in particular for detailed segmentations it will require staff with neuroanatomical expertise and replication of the method between different studies can be difficult. [22, 23] One previous study [24] have performed a semi-automated segmentation, relying on initial manual identification of the whole amygdala and hippocampus, and one study [25] have performed a manual segmentation of hippocampus into two subregions. Kim et al. [24] reported smaller hippocampal CA1 and smaller laterobasal and centromedial nuclear amygdala subdivisions in NT1 patients (n=33) compared to controls (n=31) [24] and Křečková et al. [25] reported smaller anterior hippocampal volume in NT1 patients (n=48) compared to healthy controls (n=37).

Possibly reflecting gliosis due to neuroinflammation or processes related to the increased numbers of histaminergic neurons in NT1, we have previously conducted a hypothalamic subregion segmentation in the same 54 NT1 patients versus 114 healthy controls, and found larger volume of bilateral tubular-inferior hypothalamic subregions in NT1 patients compared to controls [26]. Furthermore, in the same sample we found no significant case-control differences in 10 MRI-based subcortical total volume measures, including bilateral thalamus, amygdala, and hippocampus [27]. However, recognizing the potential for subregional differences, we here aim to further explore these critical regions using recent methods for MRI-based subregion segmentations.

To this end, we performed an automated subregion segmentation of the hippocampus, amygdala, thalamus and brainstem [22, 23, 28–31] using FreeSurfer [32]. We performed case-control comparisons using general linear models. To the best of our knowledge this is the first study to perform an MRI-based automated segmentation of the hippocampus and amygdala subregions comparing NT1 patients and healthy controls that does not rely on any manual delineation. As far as we know there has not previously been performed any type of MRI-based thalamus and brainstem segmentation comparing NT1 patients and healthy controls.

## 2. Materials and Methods

### 2.1 Participants

We recruited 70 suspected NT1 patients with disease onset after 2009/2010 H1N1-vaccination campaign, in a national follow-up project initiated by the Norwegian Ministry of Health and Care Services, from June 2015 to April 2017. The Norwegian Centre of Expertise for Neurodevelopmental Disorders and Hypersomnias (NevSom) held narcolepsy disease education and counseling courses for patients with their families, and in relation with this they were additionally invited to participate in a research project conducted while staying at the center.

The study was approved by the Norwegian regional committees for medical and health research ethics (REK) and written informed consent was obtained from all participants. For children <16 years of age parents would sign. Exclusion criteria for patients were severe psychiatric, somatic or neurological disorders, neuroradiological findings requiring a clinical follow-up, metallic implants and previous head injury with loss of consciousness for 10 minutes or 30 minutes amnesia. We excluded two patients due to severe neurological/psychiatric disorders, 12 patients were excluded due to MRI-data of insufficient quality either due to dental braces or motion artifacts, and two patients were excluded as they did not fulfill the ICSD-3 diagnostic criteria for NT1 [33].

Table 1 summarizes key clinical and demographic variables of the included patients and healthy controls. Hypocretin levels had been measured inn 51 patients that all were hypocretin-deficient (CSF-hypocretin-1 level < 1/3 of the normal mean). The three patients that had not measured CSF-hypocretin were all HLA-DQB1*0602-positive and had typical cataplexy.

**Table 1.**
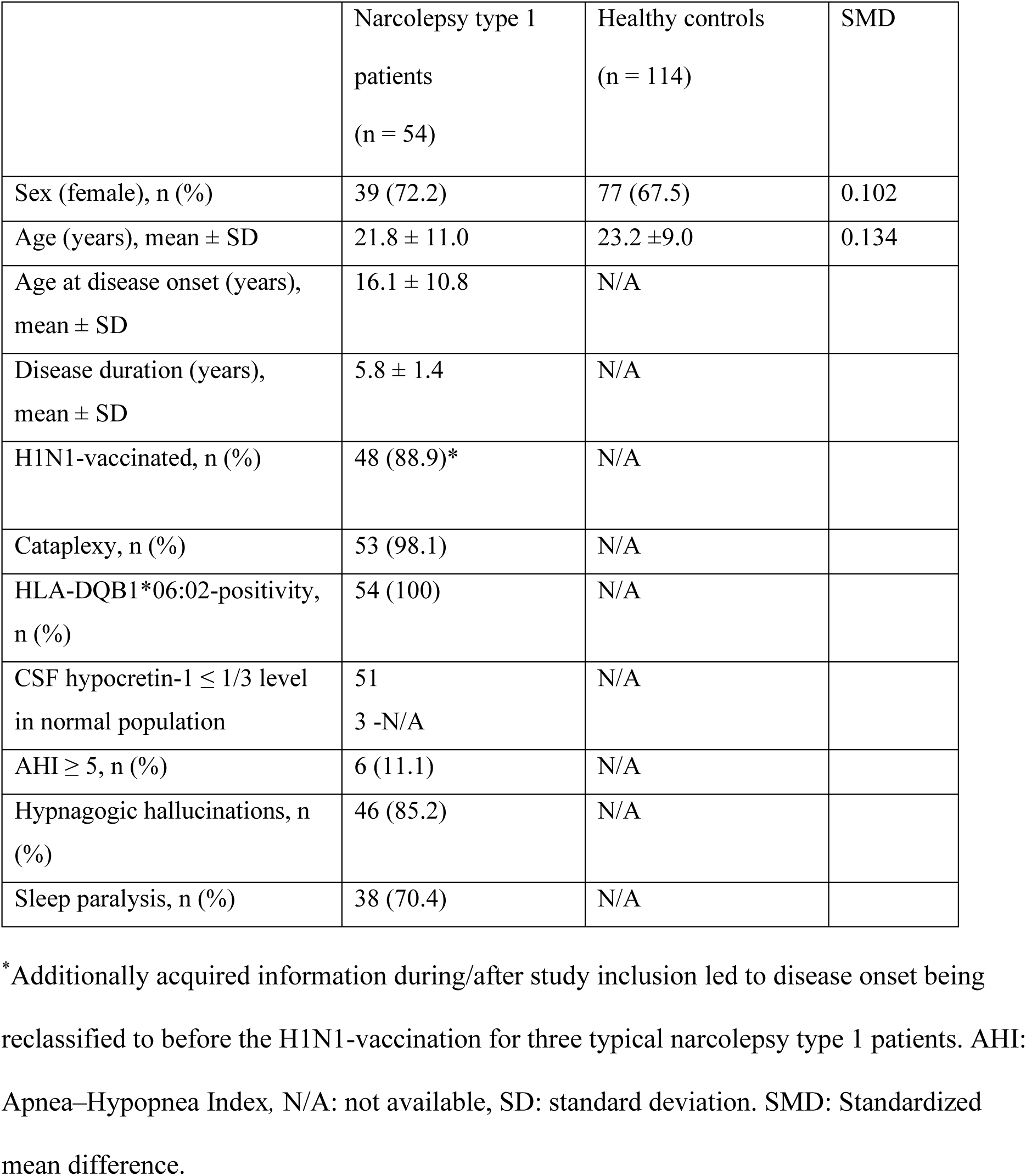
Demographic and clinical data.

Patients were requested to stop any medication affecting sleep/wake including narcolepsy medication for 14 days prior to inclusion. Three patients were not able to stop medication that could influence sleep/wake, one patient only paused Venlafaxine for a week prior to inclusion due to severe cataplexy, one patient continued to use Catapresan and another patient continued to use Fluoxetine for comorbidities.

During or after the inclusion it was discovered that three patients had disease onset prior to the H1N1-vaccination. They were typical NT1 patients with hypocretin-deficiency, cataplexy and HLA-DQB1*06:02-positivity and were therefore kept in the study.

The information about the H1N1-vaccination in Norway was obtained from the official Norwegian Immunisation Registry (SYSVAK). Pandemrix® was the main vaccine used in Norway for H1N1 [34], however for those with severe egg allergy Celvapan was an option [35]. There were two patients that reported being H1N1-vaccinated in their workplace but were not registered in SYSVAK, these patients were counted in the H1N1-vaccinated group.

Since comorbidities are frequent in NT1 patients [5] we included the following: Asperger syndrome (n=2), attention-deficit hyperactivity disorder (ADHD; n=1), migraine (n=6), anxiety (n=1), depression (n=1), hypothyroidism (n=1), Tourette syndrome (n=1), kidney disease (n=1), type 2 diabetes (n=1) and premature birth without severe long-term complications (n=1). As some NT1 patients had more than one comorbidity there were in total 14 patients with comorbidity.

Healthy controls (n=114) were included from ongoing collaborating studies using the same MRI-scanner and protocol. Healthy controls were excluded if they had somatic, neurological or psychiatric disease, metal implants, severe psychiatric family history, previous head injury with 30 minutes amnesia or loss of consciousness for 10 minutes or neuroradiological findings indicating previous or ongoing disease or abnormalities.

We have previously reported on the same 54 patients and 114 controls in a brain-wide T1-weighted MRI case-control study investigating cortical thickness and sub-cortical volumes [27] and in a T1-weighted MRI study focusing on the segmentation of the hypothalamus [26]. Additionally, 51 of the patients have been included in a MRI DTI study [36] and 37 in a fMRI study [37] obtained in the same session.

### 2.2 Narcolepsy diagnosis

NT1 diagnosis was verified according to ICSD-3 criteria [33] by an experienced neurologist and somnologist (S.K.H). The protocol included semi-structured interviews and questionnaires, including the Stanford Sleep Questionnaire and Epworth Sleepiness scale [38] translated to Norwegian, in addition to a neurological examination, routine blood parameter sampling, actigraphy, human leukocyte antigen (HLA)-typing, polysomnography (PSG) and a multiple sleep latency test (MSLT). Spinal taps for the patients were done prior to or after inclusion at their local hospital. As previously reported, CSF hypocretin-1 levels were analyzed using a modified version of the Phoenix Pharm. Inc. radioimmunoassay methods [39, 40].

### 2.3 Polysomnography recordings

Prior to the PSG an actigraphy recording (Philips Actiwatch, Respironics Inc., Murrysville, PA, USA) was obtained; 34 patients were studied for 12-14 days (one patient not consistently), 14 patients for 8-11 days (three patients not consistently), two patients wore the actigraphy very inconsistently, and 5 patients were without actigraphy. The SOMNOmedics system (SOMNOmedics GmbH, Randersacker, Germany) were used for PSG recordings with the electrodes F4-A1, C4-A1, O2-A1, F3-A2, C3-A2, and O1-A2, surface electromyography of the tibialis anterior and submentalis muscles, vertical and horizontal electro-oculography, nasal thermistor, nasal pressure transducer, thoracic respiratory effort, oxygen saturation and electrocardiography. Preferably impedance was kept at 5Ω, but below 10kΩ was also accepted. The MSLT was performed the day after the PSG with 5-naps (30 minutes) which each were separated by 2 hours. The American Academy of Sleep Medicine scoring criteria were used [33].

### 2.4 MRI acquisition and processing

T1-weighted-3D-BRAVO MRI data was obtained with a General Electric Discovery MR750 3T scanner with a 32-channel head coil with the following parameters: echo time (TE): 3.18 ms; repetition time (TR): 8.16 ms; Voxel size 1 x 1 x 1 mm; flip angle: 12°; 188 sagittal slices. The T1-weighted data was processed with FreeSurfer [32] (v.7.1.0) using the standard recon-all pipeline. The hippocampal and amygdala segmentation [22, 23] was performed with a tool in FreeSurfer, which have been developed by utilizing ex vivo MRI data of ultra-high resolution to create a probabilistic atlas through an Bayesian inference atlas building algorithm [22, 23, 41]. We obtained nine amygdala nuclei (central, cortical, medial, lateral, basal, accessory basal, anterior amygdaloid, cortico-amygdaloid transition area, paralaminar nuclei) and the whole amygdala volume for the left and right side [23, 41]. An overview of the hippocampal subregions obtained from the segmentation can be found in Table 2. Additionally, with tools from FreeSurfer, we performed thalamus segmentations using Bayesian inference based on a probabilistic atlas built with histological data and ex vivo and in vivo brain MRI scans [28, 29] and brainstem segmentations using a Bayesian algorithm relying on a probabilistic atlas [30, 31]. An overview of the thalamic subregions can be found in Table 4. The brainstem was segmented into the medulla oblongata, midbrain, pons and superior cerebellar peduncle (SCP).

**Table 2.**
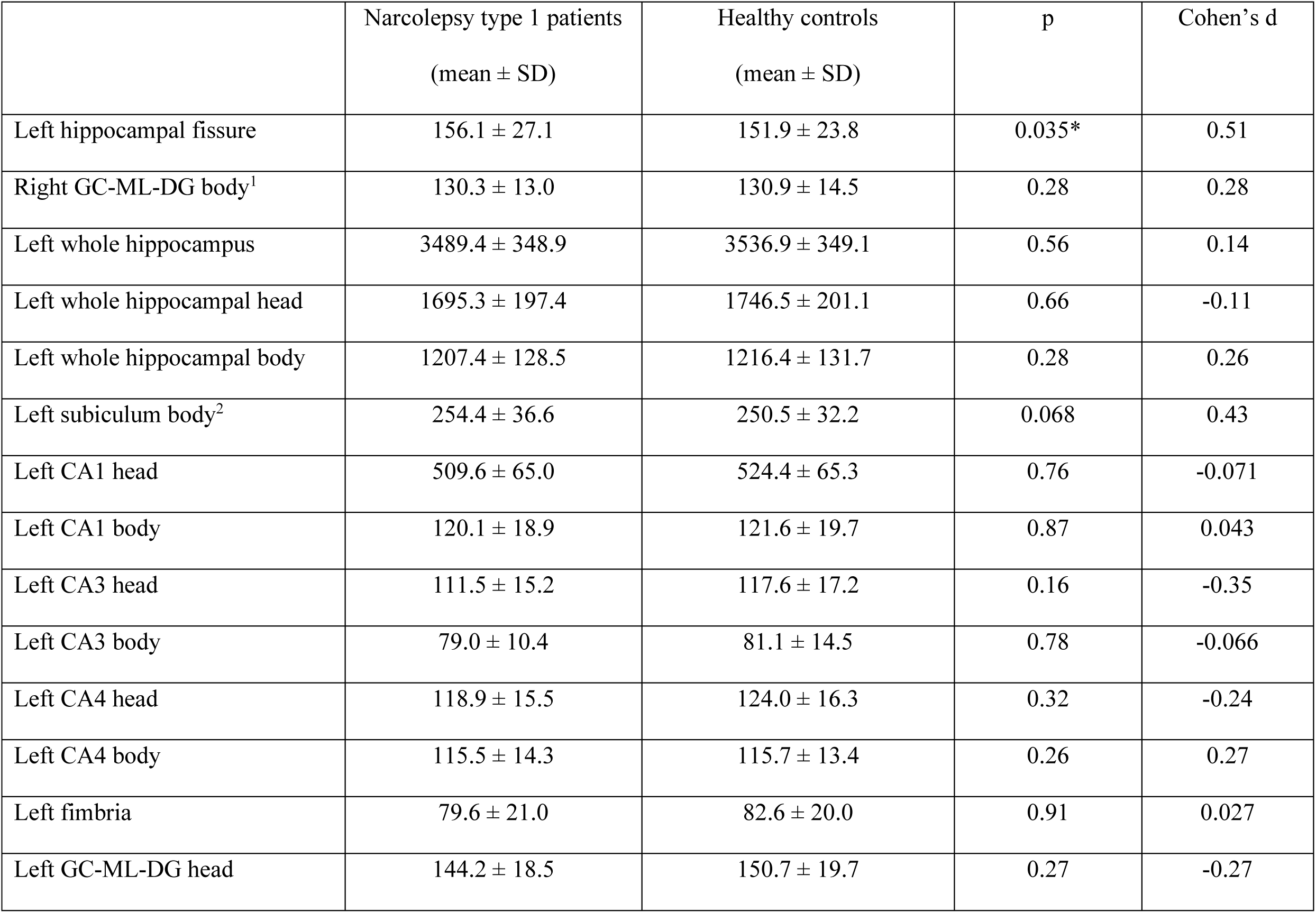
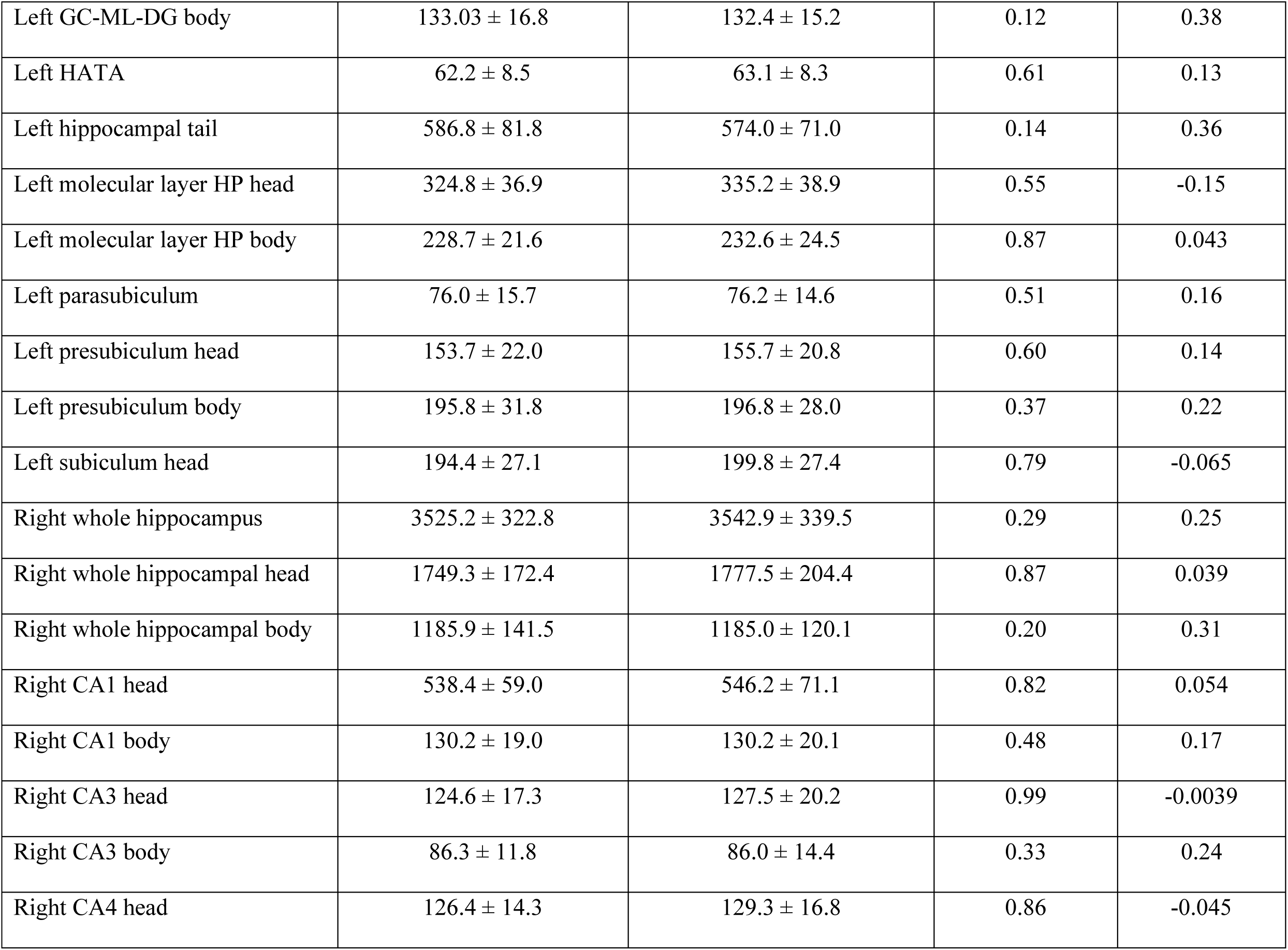
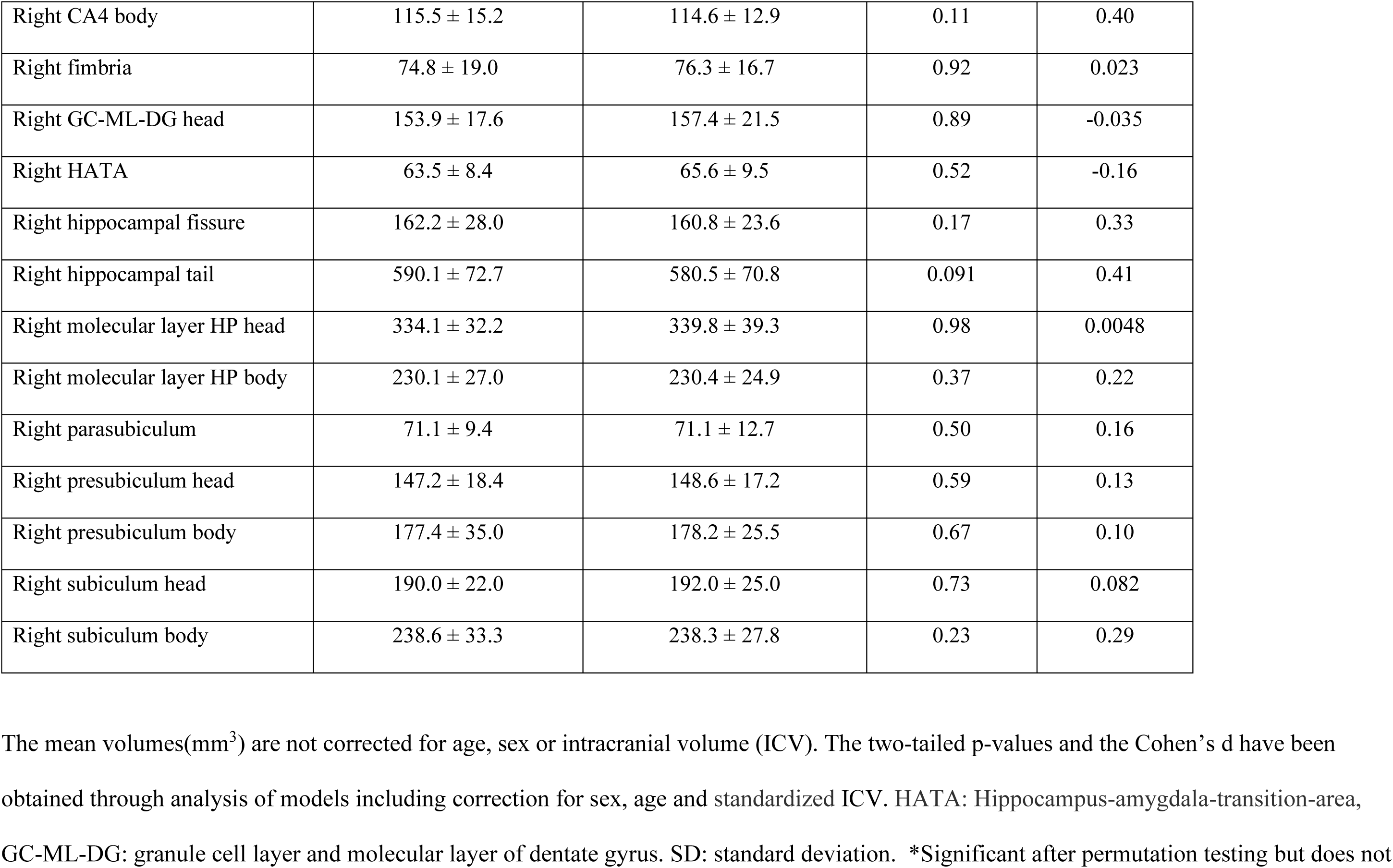
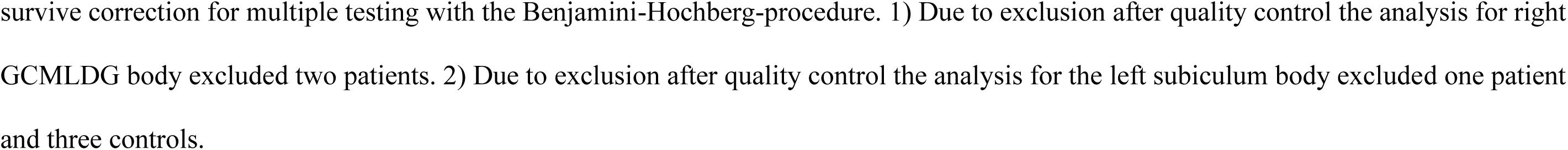
Uncorrected mean volumes for the hippocampus segmentation for narcolepsy type 1 patients and controls.

### 2.5 Statistical analysis

Quality control was conducted by evaluating the effects of outliers. A preliminary multiple regression analysis was performed in R [42], including sex, age, and intracranial volume (ICV) as independent variables, while treating the various subregions from the segmentations as dependent variables. Outliers in the residuals were defined as values 1.5 times the interquartile range (IQR) above the third quartile and below the first quartile. A subsequent multiple regression analysis was conducted, excluding these outliers. The results from the analyses with and without outliers were compared. If the presence of outliers influenced the results - causing a change from significant to non-significant group differences or vice versa-these outliers were visually inspected and considered for exclusion.

We performed the main analysis in Permutation Analysis of Linear Models (PALM) [43, 44] (https://web.mit.edu/fsl_v5.0.10/fsl/doc/wiki/PALM.html), where we tested for group differences with general linear models and permutation testing. Following segmentation, we obtained 44 subregions for the hippocampus (including whole left and right hippocampus regions), 20 subregions for the amygdala (including whole left and right amygdala regions), 52 subregions for the thalamus (including whole left and right thalamus regions), and five subregions for the brainstem (including the whole brainstem). In the main analysis we included age, sex and ICV and performed 5000 permutations. Outputs from the analysis were t-statistics, two-tailed p-values, contrast of parameter estimate (COPE)s and Cohen’s ds. The definition of Cohen’s d in PALM is COPE / square root (variance of the residuals). Exchangeability was not constrained except for a sibling pair in the patient group where it had to be constrained to account for the lack of independence. The Benjamini-Hochberg procedure [45] were used to control the false discovery rate (FDR) at 5 %.

## 2.6 Data availability

The data are not publicly available due to privacy concerns and restrictions imposed by our ethics approval. Given that Norway has a relatively small population and NT1 is a rare disorder, the combination of variables may therefore lead to the identification of individuals.

## 3. Results

### 3.1 Hippocampus subregions

Figure 1 shows a single-subject hippocampus and amygdala segmentation that has been overlaid onto a standard MNI 152 T1-image. Table 2 summarizes the mean uncorrected volumes as well as the stats and effect sizes from the case-control comparisons (corrected for age, sex and ICV) for the hippocampus subregions. Permutation testing revealed no case-control differences that remained significant after FDR correction. Due to exclusion after quality control the analysis for right granule cell layer and molecular layer of dentate gyrus (GC-ML-DG) body excluded two patients and the analysis for the left subiculum body excluded one patient and three controls.

**Figure 1.**
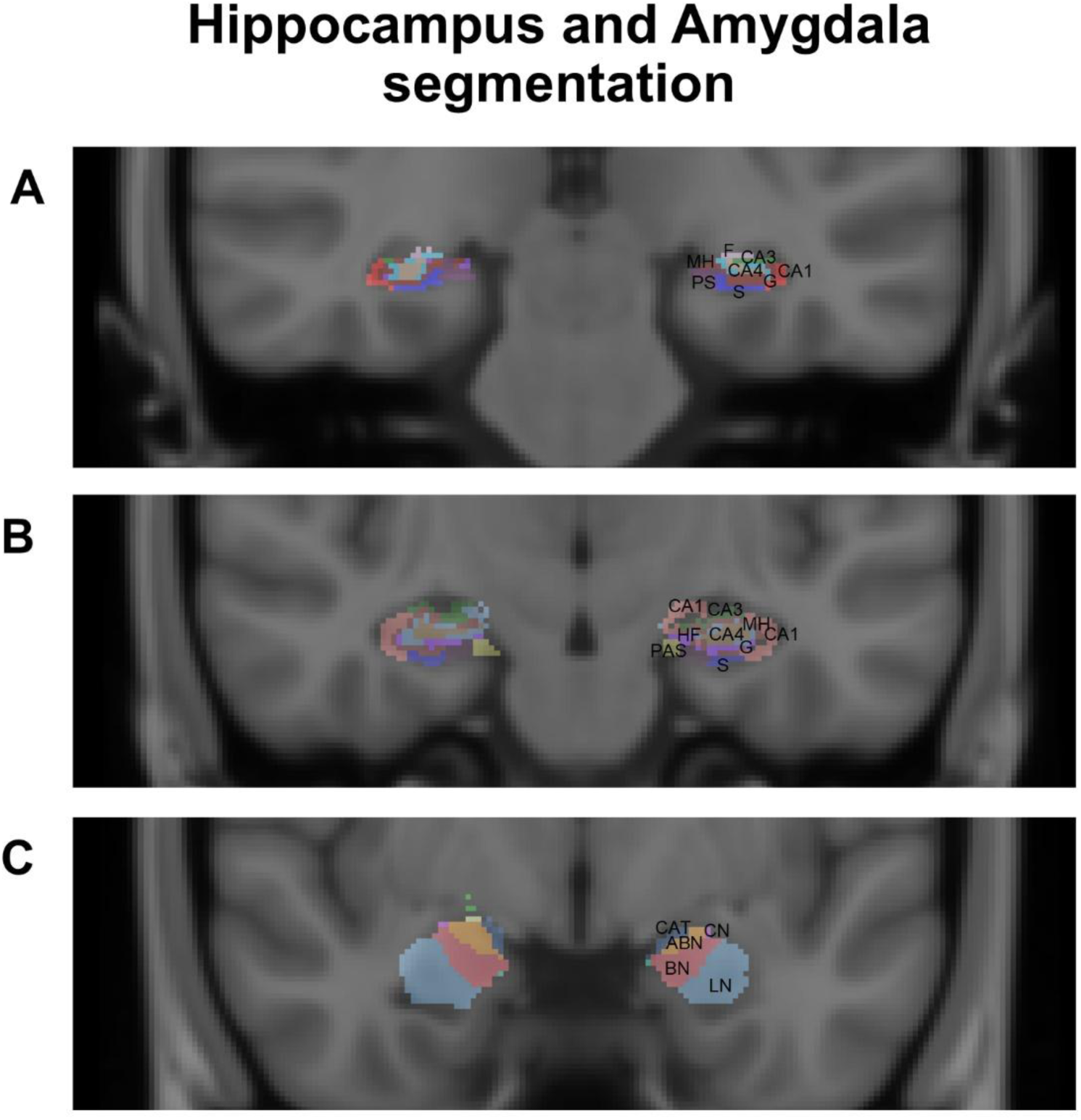
Example of Hippocampus and Amygdala segmentation. A single-subject hippocampus and amygdala segmentation has been overlaid onto a standard MNI 152 T1-image. The figure shows some of the subregions from the hippocampus and amygdala segmentation, for a full overview of the subregions in the amygdala segmentation see Table 3 for the hippocampus segmentation see Table 2. **A)** red = CA1-body, blue = subiculum-body (S), green = CA3-body, beige = CA4-body, turquoise = GC-ML-DG (granule cell layer and molecular layer of dentate gyrus) -body (G), pink = fimbria (F), brown = molecular layer HP-body (MH), purple = presubiculum-body (PS), **B)** beige = CA4-head, turquoise = GC-ML-DG (granule cell layer and molecular layer of dentate gyrus) -head (G), purple = hippocampal fissure (HF), blue = subiculum-head (S), dark yellow = parasubiculum (PAS), brown = molecular layer HP-head (MH), dark pink = CA1-head, green = CA3-head, **C)** light blue = lateral nucleus (LN), red = basal nucleus (BN), orange = accessory basal nucleus (ABN), purple = central nucleus (CN), dark blue = cortico-amygdaloid-transition area (CAT).

**Table 3.** Uncorrected mean volumes for the amygdala segmentation for narcolepsy type 1 patients and controls.

| | Narcolepsy type 1<br>patients<br>(mean ± SD) | Healthy controls<br>(mean ± SD) | p | Cohen's d |
| --- | --- | --- | --- | --- |
| Left lateral nucleus | 624.8 ± 66.2 | 655.7 ± 66.5 | 0.021* | -0.54 |
| Left medial nucleus | 21.2 ± 5.7 | 19.5 ± 5.3 | 0.040* | 0.50 |
| Right lateral nucleus | 653.2 ± 66.4 | 683.0 ± 64.6 | 0.032* | -0.51 |
| Left whole amygdala | 1679.3 ± 174.1 | 1741.6 ± 164.9 | 0.13 | -0.36 |
| Left accessory basal<br>nucleus | 250.8 ± 28.4 | 258.8 ± 27.3 | 0.34 | -0.23 |
| Left anterior amygdaloid<br>area | 51.6 ± 6.0 | 53.9 ± 6.8 | 0.082 | -0.41 |
| Left basal nucleus | 434.3 ± 49.9 | 452.1 ± 46.4 | 0.11 | -0.38 |
| Left central nucleus | 43.1 ± 9.0 | 42.4 ± 6.8 | 0.26 | 0.27 |
| Left cortical nucleus | 23.3 ± 3.8 | 23.6 ± 3.8 | 0.94 | -0.018 |
| Left cortico-amygdaloid<br>transition area | 178.7 ± 20.5 | 183.0 ± 20.3 | 0.73 | -0.084 |
| Left paralamina nucleus | 51.5 ± 6.6 | 52.6 ± 6.4 | 0.98 | -0.0057 |
| Right whole amygdala | 1773.1 ± 172.6 | 1835.0 ± 168.6 | 0.14 | -0.34 |
| Right accessory basal<br>nucleus | 270.9 ± 29.8 | 277.9 ± 29.0 | 0.54 | -0.14 |
| Right anterior<br>amygdaloid area | 58.4 ± 6.2 | 60.3 ± 7.0 | 0.27 | -0.26 |
| Right basal nucleus | 455.9 ± 48.2 | 474.1 ± 47.0 | 0.090 | -0.41 |
| Right central nucleus<sup>1</sup> | 48.6 ± 10.1 | 47.4 ± 8.1 | 0.057 | 0.45 |
| Right cortical nucleus | 25.8 ± 4.4 | 26.3 ± 4.0 | 0.96 | -0.013 |
| Right cortico-amygdaloid transition area | 185.8 ± 20.0 | 191.4 ± 20.3 | 0.34 | -0.22 |
| Right medial nucleus | 22.9 ± 7.4 | 21.9 ± 6.0 | 0.17 | 0.32 |
| Right paralamina nucleus | 51.1 ± 5.7 | 52.8 ± 5.7 | 0.28 | -0.26 |

The mean volumes(mm<sup>3</sup>) are not corrected for age, sex or intracranial volume (ICV). The two-tailed p-values and the Cohen's d have been obtained through analysis of models including correction for sex, age and standardized ICV. SD: standard deviation \*Significant after permutation testing but does not survive correction for multiple testing with the Benjamini-Hochberg-procedure. 1) Due to exclusion after quality control the analysis for right central nucleus excluded one patient.

### 3.2 Amygdala subregions

Table 3 summarizes the mean uncorrected volumes as well as stats and effect sizes from the case-control comparisons (corrected for age, sex and ICV) for the amygdala subregions. Permutation testing revealed no case-control differences that remained significant after FDR correction. Due to exclusion after quality control the analysis for right central nucleus excluded one patient.

### 3.3 Thalamus subregions

Figure 2 shows a single-subject thalamus segmentation that has been overlaid onto a standard MNI 152 T1-image. Table 4 summarizes the mean uncorrected volumes as well as the stats and effect sizes from case-control comparisons (corrected for age, sex and ICV) for the thalamus subregions. Permutation testing revealed no case-control differences that remained significant after FDR correction. Due to exclusion after quality control the analysis for left lateral geniculate (LGN) excluded one control, the analysis for the left pulvinar inferior (PuI) excluded one control and the analysis for left pulvinar medial (PuM) excluded one patient and three controls.

**Table 4.**
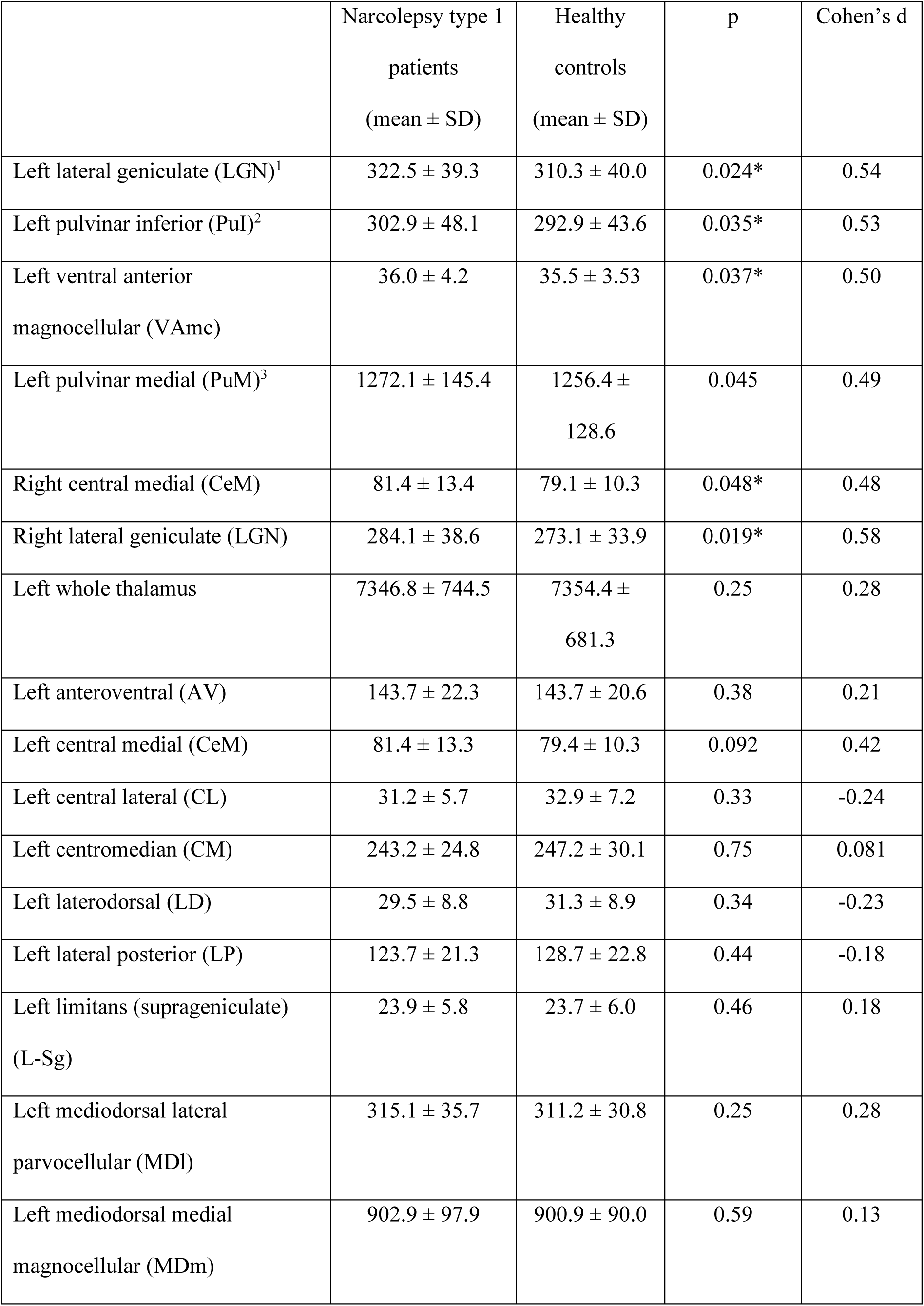
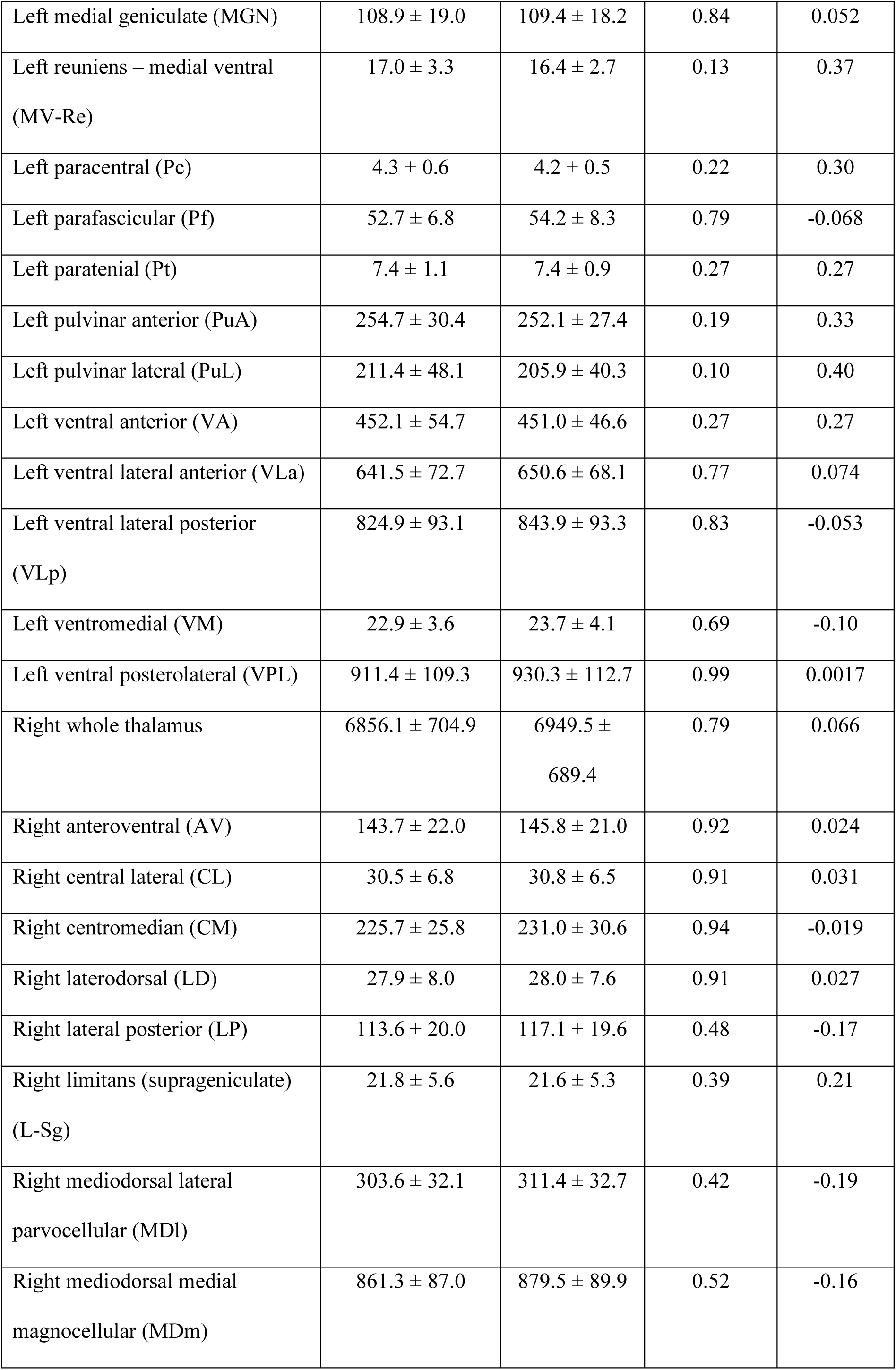
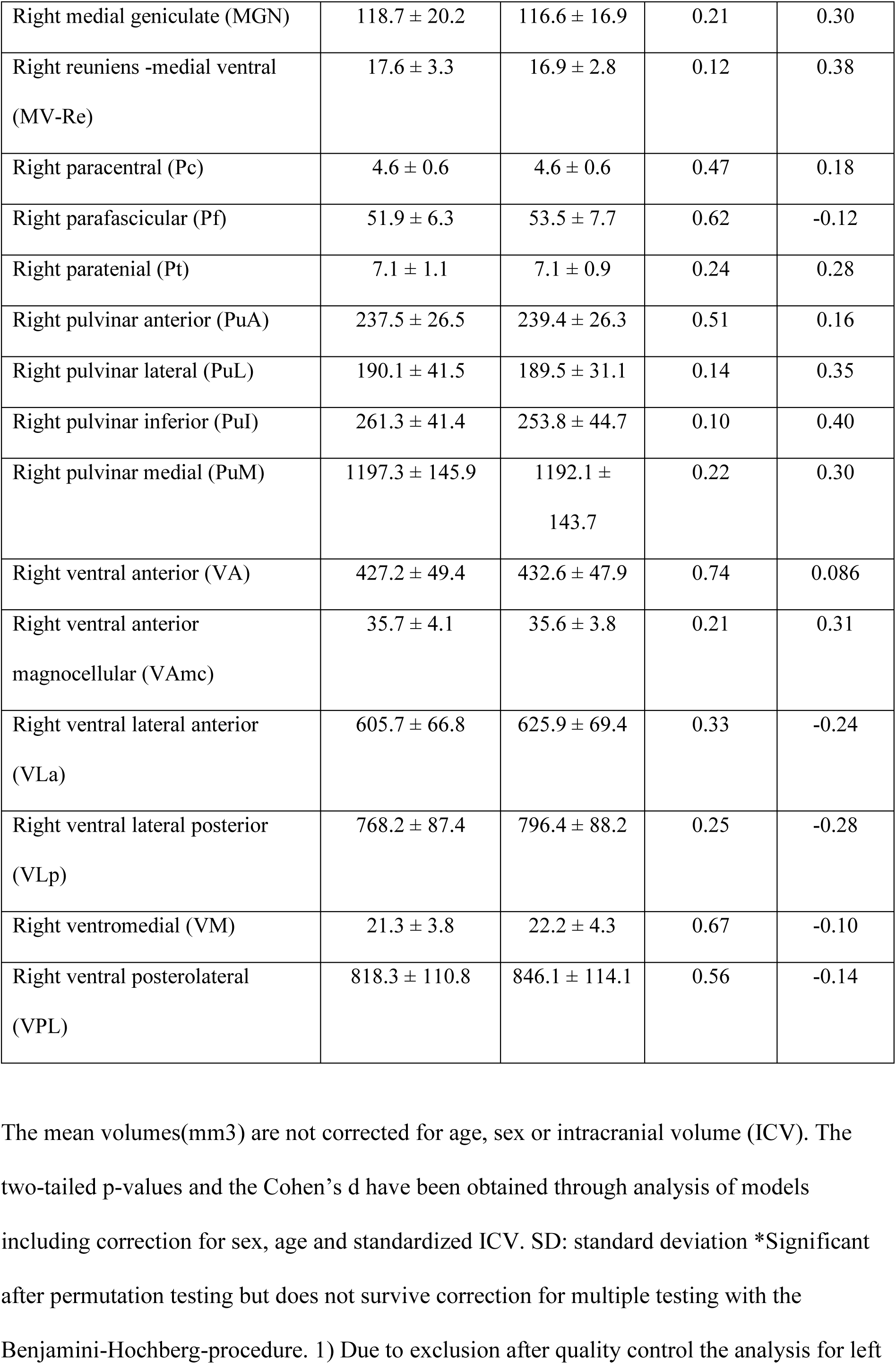
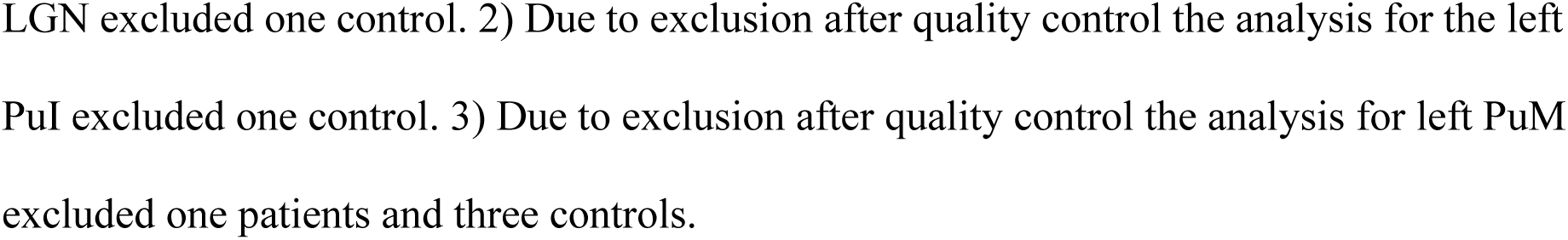
Uncorrected mean volumes for the thalamus segmentation for narcolepsy type 1 patients and controls.

**Figure 2.**
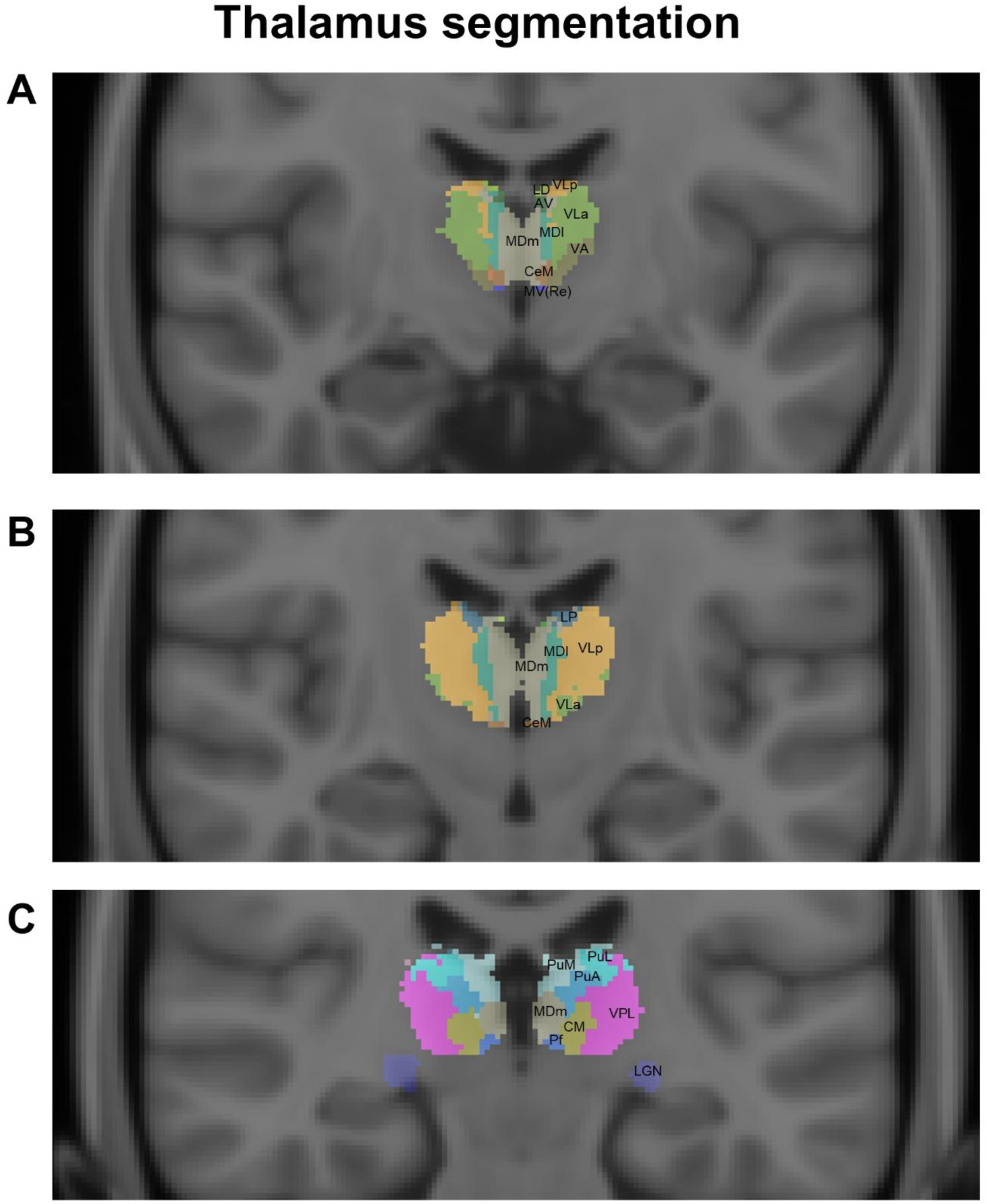
Example of Thalamus segmentation. A single-subject thalamus segmentation that has been overlaid onto a standard MNI 152 T1-image. The figure shows some of the subregions from the thalamus segmentation, for a full overview of the thalamus segmentation see Table 4, **A)** light green (lateral) = ventral lateral anterior (VLa), yellow = ventral lateral posterior (VLp), light green (superior) = laterodorsal (LD), green= anteroventral (AV), bluish green (medial) = mediodorsal lateral parvocellular (MDl), beige (medial) = mediodorsal medial magnocellular (MDm), brown = central medial (CeM), brownish green = ventral anterior (VA), blue = Reuniens – medial ventral (MV(Re)) **B)** light green (lateral) = ventral lateral anterior (VLa), yellow = ventral lateral posterior (VLp), bluish green (medial) = mediodorsal lateral parvocellular (MDl), beige = mediodorsal medial magnocellular (MDm), brown = central medial (CeM), blue = lateral posterior (LP) **C)** purple = ventral posterolateral (VPL), yellowish brown = centromedian (CM), beige = mediodorsal medial magnocellular (MDm), turquoise = pulvinar lateral (PuL), blue (next to purple) = pulvinar anterior (PuA), light blue = pulvinar medial (PuM), blue = parafascicular (Pf), dark blue (inferior) = lateral geniculate (LGN)

### 3.4 Brainstem subregions

Figure 3 shows a single-subject brainstem segmentation that has been overlaid onto a standard MNI 152 T1-image. Table 5 summarizes the mean uncorrected volumes as well as the stats and effect sizes from the case-control comparisons (corrected for age, sex and ICV) for the brainstem subregions. Permutation testing revealed no case-control differences that remained significant after FDR correction.

**Figure 3.**
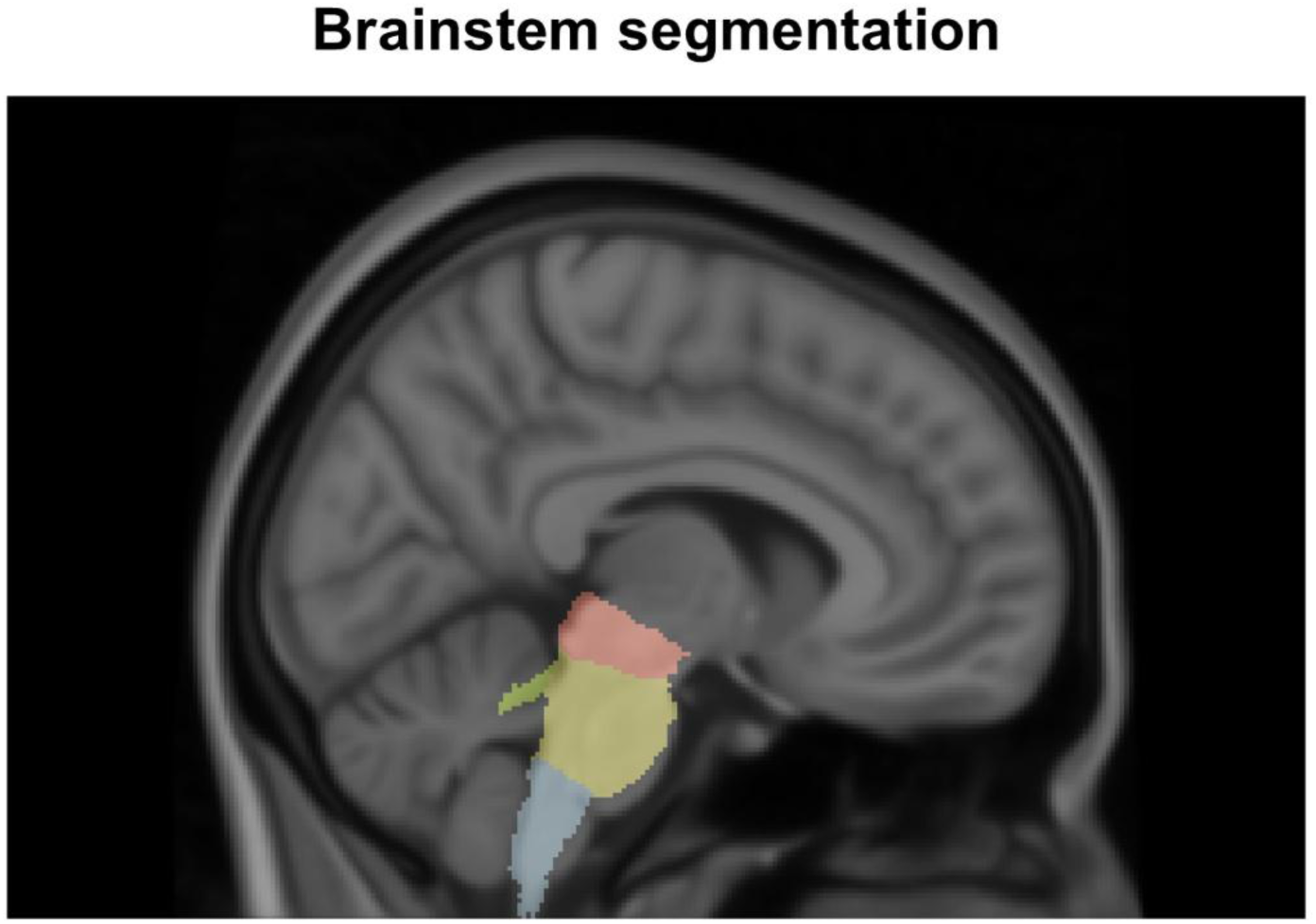
Example of Brainstem segmentation. A single-subject brainstem segmentation that has been overlaid onto a standard MNI 152 T1-image. Light blue = medulla oblongata, yellow = pons, red = midbrain, green = superior cerebellar peduncle (SCP)

**Table 5.** Uncorrected mean volumes for the brainstem segmentation for narcolepsy type 1 patients and controls.

| | Narcolepsy type 1 patients<br>(mean ± SD) | Healthy controls<br>(mean ± SD) | p | Cohen's d |
| --- | --- | --- | --- | --- |
| Whole brainstem | 25283.7 ± 2631.3 | 25326.3 ± 3119.9 | 0.057 | 0.47 |
| Medulla oblongata | 4524.8 ± 463.2 | 4523.5 ± 573.8 | 0.078 | 0.45 |
| Midbrain | 6063.6 ± 581.7 | 6084.8 ± 668.9 | 0.10 | 0.40 |
| Pons | 14450.6 ± 1657.5 | 14471.9 ± 1975.7 | 0.069 | 0.44 |
| SCP | 244.8 ± 34.2 | 246.2 ± 44.3 | 0.28 | 0.27 |

The mean volumes(mm3) are not corrected for age, sex or intracranial volume (ICV). The two-tailed p-values and the Cohen's d have been obtained through analysis of models including correction for sex, age and standardized ICV. SD: standard deviation, SCP: superior cerebellar peduncle.

## 4. Discussion

Previous studies have implicated the amygdala, thalamus, brainstem, and hippocampus in the brain networks involved in sleep-wake regulation and NT1, with mixed and inconclusive results. Most previous human imaging studies comparing subcortical brain volumes between patients with NT1 and controls have not considered subregional volumes, which have just recently become accessible using fully automated MRI based methods.

The brainstem and the thalamus have important roles in sleep-wake regulation, for example the parabrachial nucleus and the pedunculopontine tegmental nucleus in the pons are considered key parts of the arousal system [7]. Amygdala has been implicated in the brain networks involved in cataplexy, and lesions in the amygdala markedly reduced cataplexy events associated with positive emotions and high arousal in hypocretin-knockout mice [2].

Further, in narcoleptic dogs cataplexy-active neurons have been identified in the amygdala [2]. Previous studies have given conflicting results in regards to memory difficulties in NT1 patients, but impaired memory consolidation for both visual discrimination and motor skills in NT1 patients has been suggested [10]. Our analysis revealed no robust group differences between NT1 patients and controls for any of the included subregions of the hippocampus, amygdala, thalamus or the brainstem.

Most previous studies [11–13, 15, 20, 46, 47] assessing subcortical volumetry in NT1 have considered total but not subregional measures, including a study from our own group that revealed no significant group differences in the hippocampus, thalamus or amygdala total volumes [27]. Using the same sample as in the current study, we conducted an automated hypothalamic segmentation and found larger hypothalamic volume, in particularly in the left and right tubular-inferior hypothalamic subregions, in NT1 patients compared to controls [26], conceivably reflecting gliosis or processes linked to increased number of histaminergic neurons.

One previous study [24] investigating amygdala and hippocampus subregions semi-automated in 33 drug-naïve NT1 patients and 31 age-/sex-matched controls (mean ± SD: 27±6 years) reported smaller amygdala laterobasal and centromedial nuclear subdivisions and smaller hippocampal CA1 volume in narcolepsy patients compared to controls. Using manual segmentation of brain scans from 48 patients with narcolepsy and 37 controls, Křečková et al. reported volume loss in the anterior parts of the hippocampus in the patient group [25].

Our current results differ from Kim et al. [24], which could be due to difference in segmentation methods or patient characteristics and sample size. Several factors may account for the discrepancies between our current findings and the results reported by Křečková et al. [25], including patient characteristics such as disease duration (18.6 ±14.8 years vs 5.8 ± 1.4 years) and MRI field strength (1.5 Tesla vs 3 Tesla). Křečková et al. [25] also acknowledge the age heterogeneity of their patient sample as one of their main limitations. The patient and control population in Křečková et al.2019 [25] overlapped with samples in Nemcova et al. 2015 [15] and Brabec et al. 2011 [11]. The smaller Brabec et al. [11] study reported smaller volume of bilateral amygdala in 11 patients compared to 11 controls.

Nemcova et al. [15] reported no significant group differences in amygdala volume between narcolepsy patients with cataplexy, narcolepsy patients without cataplexy and controls. However, the volume of hippocampus was significantly smaller in the narcolepsy patients with cataplexy compared to the two other groups. Jo et al. 2012 [20] found smaller bilateral hippocampal volume in patients (n=36) compared to controls (n=36), but no correlation between hippocampal volume and memory scores in the patient group. Tondelli et al. 2018 [21] reported larger total volume in the right hippocampus among 20 drug-naïve children/adolescents with NT1 compared to 19 healthy controls.

We are not aware of previous MRI studies using automated thalamus segmentation in NT1 patients. Using voxel-based morphometry (VBM), Joo et al. 2009 [12] reported lower gray matter concentration in thalamus bilaterally in patients compared to controls, and no significant case-control differences in hippocampus and amygdala. Similarly, Kim et al. 2009 [14] reported that NT1 patients had smaller total gray matter volume in the left thalamus and the brainstem compared to controls, but not in the hippocampus and amygdala. Several previous MRI-studies [13, 16–19] reported no significant case-control differences in the volumes of hippocampus, thalamus and amygdala. Kaufmann et al. [13] reported no significant case-control differences in amygdala. Draganski et al. [16], Overeem et al. [17], Scherfler et al. [19] and Schaer et al. [18] reported no significant case-control differences in amygdala, hippocampus, or thalamus total volume.

Our analysis revealed no robust volume differences between patients and controls in the abovementioned regions. In addition to small sample size in some of the previous studies, which increases the probability both for Type I and Type II errors, any divergent findings between the current and previous studies might be partly attributed to patient sample characteristics. For example, our NT1 patient sample is known hypocretin deficient i.e. highly homogeneous, while the hypocretin status was unknown/not reported in the other segmentation studies. [24, 25]

Moreover, the patients in our current study were post-H1N1(largely Pandemrix®-vaccinated) individuals. However, although some studies have reported possible differences like a more sudden onset of symptoms [3, 4, 48–53], most studies point to sporadic narcolepsy and narcolepsy linked to the Pandemrix®-vaccination being the same phenotype [3, 53, 54].

As previously mentioned, an earlier MRI study conducted by our group [26] involved hypothalamic automated segmentation in the same sample of patients and controls as used in the current study. The results indicated that patients exhibited significantly larger hypothalamic volumes, particularly in the tubular-inferior subregions, compared to controls. This observation of increased hypothalamic volume may be specific to post-H1N1 narcolepsy. However, it is important to note that MRI-based hypothalamic segmentation has not yet been performed on a large, sporadic NT1 sample.

Additionally, another comprehensive MRI study by our group [27] assessed group differences between NT1 patients and controls across brain-wide subcortical volume measures and cortical thickness measures. This study revealed that NT1 patients had a thinner entorhinal cortex, superior temporal gyrus, and temporal poles compared to the control group. The divergence in findings may suggest that these characteristics are specific to post-H1N1 narcolepsy. Alternatively, the differences could be attributed to the increased statistical power in our study [27], which included nearly twice the number of patients compared to earlier studies.

The current automated subregional segmentation algorithms have limitations in regards to their interpretations as the internal boundaries between the substructures relies largely on prior knowledge which is summarized in the statistical atlases [41]. For example for hippocampus volumes of internal subregions like granule cell layer of dentate gyrus (GC-DG), CA4 and molecular layer, must be interpretated with caution while the fimbria and the tail are more reliable [41].

To conclude, our analysis revealed no robust case-control volume differences between NT1 patients and healthy controls in any of the included subregions of the hippocampus, thalamus, amygdala and brainstem. In the future we will need large multi-site studies to gain sufficient power to have the possibility to detect even more subtle group differences.

## 5. Acknowledgments

A special thank you goes to the participating patients and controls. We also want to thank Janita Vevelstad (nurse at the Unit for Brain Disorders, Department of Rare Diseases, Oslo University Hospital) and Rannveig Viste (PhD, nurse and molecular biologist at the Unit for Brain Disorders, Department of Rare Diseases, Oslo University Hospital) for being a part of the data collection and sleep scoring of polysomnography/multiple sleep latency tests. Furthermore, we would like to thank Ranveig Østrem’s (bioengineer at the Hormone Laboratory at Oslo University Hospital) for her work with biobanking and -analyses and Marte K. Viken (PhD, senior researcher at the Oslo University Hospital and University of Oslo) for her work with the HLA sequencing.

## Funding

S. K. H was partially funded by research support from the Norwegian Ministry of Health and Care Services. H.T.J was during data collection fully funded by the Norwegian Ministry of Health and Care Services and during data analyses and manuscript writing partly by the Helse SØ grant (2019032) and partly by the Norwegian Ministry of Health and Care Services. D. A was supported by the South-Eastern Norway Regional Health Authorities (2019107, 2020086). The European Union’s Horizon 2020 Research and Innovation program (ERC StG, Grant # 802998), the Research Council of Norway (223273, 249795, 300767) and the South-Eastern Norway Regional Health Authority (2019101) was also part of funding this study. The funding sources had no involvement with the study design, data collection, analysis or interpretation of the data, the writing of the article or in the decision to submit the article for publication.

## 6. Declaration of competing interests

H.T.J reports no competing interests. D.A. reports no competing interests. I.A. received speaker’s honorarium from Lundbeck. O.A.A. is a consultant for HealthLytix and has received speaker’s honorarium from Lundbeck and Sunovion. A.S. reports no competing interests. P.M.T. reports no competing interests. L.T.W reports no competing interests. S.K.H have been lecturing about narcolepsy for UCB Pharma, AOP Orphan, Jazz Pharmaceuticals, Lundbeck AS, honorarium have been paid to Norwegian Centre of Expertise for Neurodevelopmental Disorders and Hypersomnias (NevSom). S.K.H. has received honorarium for being an expert consultant for the Norwegian state regarding narcolepsy and Pandemrix.

## Notes

### Competing Interest Statement

H.T.J reports no competing interests. D.A. reports no competing interests. I.A. received speakers honorarium from Lundbeck. O.A.A. is a consultant for HealthLytix and has received speakers honorarium from Lundbeck and Sunovion. A.S. reports no competing interests. P.M.T. reports no competing interests. L.T.W reports no competing interests. S.K.H have been lecturing about narcolepsy for UCB Pharma, AOP Orphan, Jazz Pharmaceuticals, Lundbeck AS, honorarium have been paid to Norwegian Centre of Expertise for Neurodevelopmental Disorders and Hypersomnias (NevSom). S.K.H. has received honorarium for being an expert consultant for the Norwegian state regarding narcolepsy and Pandemrix.

### Author Declarations

The Norwegian Regional committees for medical and health research ethics south-east (REC) gave ethical approval for this work.

## References

1. Bassetti, C. L. A., Adamantidis, A., Burdakov, D., et al., Narcolepsy - clinical spectrum, aetiopathophysiology, diagnosis and treatment. Nat Rev Neurol, 2019. 15(9): p. 519–539.DOI: 10.1038/s41582-019-0226-9.

2. Seifinejad, A., Vassalli, A., and Tafti, M., Neurobiology of cataplexy. Sleep Med Rev, 2021. 60: p. 101546.DOI: 10.1016/j.smrv.2021.101546.

3. Sarkanen, T. O., Alakuijala, A. P. E., Dauvilliers, Y. A., and Partinen, M. M., Incidence of narcolepsy after H1N1 influenza and vaccinations: Systematic review and meta-analysis. Sleep Med Rev, 2018. 38: p. 177–186.DOI: 10.1016/j.smrv.2017.06.006.

4. Heier, M. S., Gautvik, K. M., Wannag, E., et al., Incidence of narcolepsy in Norwegian children and adolescents after vaccination against H1N1 influenza A. Sleep Med, 2013. 14(9): p. 867–71.DOI: 10.1016/j.sleep.2013.03.020.

5. Kornum, B. R., Knudsen, S., Ollila, H. M., et al., Narcolepsy. Nat Rev Dis Primers, 2017. 3: p. 16100.DOI: 10.1038/nrdp.2016.100.

6. Peyron, C., Tighe, D. K., van den Pol, A. N., et al., Neurons containing hypocretin (orexin) project to multiple neuronal systems. J Neurosci, 1998. 18(23): p. 9996–10015.

7. Saper, C. B. and Fuller, P. M., Wake-sleep circuitry: an overview. Curr Opin Neurobiol, 2017. 44: p. 186–192.DOI: 10.1016/j.conb.2017.03.021.

8. Meletti, S., Vaudano, A. E., Pizza, F., et al., The Brain Correlates of Laugh and Cataplexy in Childhood Narcolepsy. J Neurosci, 2015. 35(33): p. 11583–94.DOI: 10.1523/jneurosci.0840-15.2015.

9. Cano, C. A., Harel, B. T., and Scammell, T. E., Impaired cognition in narcolepsy: clinical and neurobiological perspectives. Sleep, 2024. 47(9).DOI: 10.1093/sleep/zsae150.

10. Cellini, N., Memory consolidation in sleep disorders. Sleep Med Rev, 2017. 35: p. 101–112.DOI: 10.1016/j.smrv.2016.09.003.

11. Brabec, J., Rulseh, A., Horinek, D., et al., Volume of the amygdala is reduced in patients with narcolepsy - a structural MRI study. Neuro Endocrinol Lett, 2011. 32(5): p. 652–6.

12. Joo, E. Y., Tae, W. S., Kim, S. T., and Hong, S. B., Gray matter concentration abnormality in brains of narcolepsy patients. Korean J Radiol, 2009. 10(6): p. 552–8.DOI: 10.3348/kjr.2009.10.6.552.

13. Kaufmann, C., Schuld, A., Pollmächer, T., and Auer, D. P., Reduced cortical gray matter in narcolepsy: preliminary findings with voxel-based morphometry. Neurology, 2002. 58(12): p. 1852–5.DOI: 10.1212/wnl.58.12.1852.

14. Kim, S. J., Lyoo, I. K., Lee, Y. S., et al., Gray matter deficits in young adults with narcolepsy. Acta Neurol Scand, 2009. 119(1): p. 61–7.DOI: 10.1111/j.1600-0404.2008.01063.x.

15. Nemcova, V., Krasensky, J., Kemlink, D., et al., Hippocampal but not amygdalar volume loss in narcolepsy with cataplexy. Neuro Endocrinol Lett, 2015. 36(7): p. 682–8.

16. Draganski, B., Geisler, P., Hajak, G., et al., Hypothalamic gray matter changes in narcoleptic patients. Nat Med, 2002. 8(11): p. 1186–8.DOI: 10.1038/nm1102-1186.

17. Overeem, S., Steens, S. C., Good, C. D., et al., Voxel-based morphometry in hypocretin-deficient narcolepsy. Sleep, 2003. 26(1): p. 44–6.

18. Schaer, M., Poryazova, R., Schwartz, S., Bassetti, C. L., and Baumann, C. R., Cortical morphometry in narcolepsy with cataplexy. J Sleep Res, 2012. 21(5): p. 487–94.DOI: 10.1111/j.1365-2869.2012.01000.x.

19. Scherfler, C., Frauscher, B., Schocke, M., et al., White and gray matter abnormalities in narcolepsy with cataplexy. Sleep, 2012. 35(3): p. 345–51.DOI: 10.5665/sleep.1692.

20. Joo, E. Y., Kim, S. H., Kim, S. T., and Hong, S. B., Hippocampal volume and memory in narcoleptics with cataplexy. Sleep Med, 2012. 13(4): p. 396–401.DOI: 10.1016/j.sleep.2011.09.017.

21. Tondelli, M., Pizza, F., Vaudano, A. E., Plazzi, G., and Meletti, S., Cortical and Subcortical Brain Changes in Children and Adolescents With Narcolepsy Type 1. Sleep, 2018. 41(2).DOI: 10.1093/sleep/zsx192.

22. Iglesias, J. E., Augustinack, J. C., Nguyen, K., et al., A computational atlas of the hippocampal formation using ex vivo, ultra-high resolution MRI: Application to adaptive segmentation of in vivo MRI. Neuroimage, 2015. 115: p. 117–37.DOI: 10.1016/j.neuroimage.2015.04.042.

23. Saygin, Z. M., Kliemann, D., Iglesias, J. E., et al., High-resolution magnetic resonance imaging reveals nuclei of the human amygdala: manual segmentation to automatic atlas. Neuroimage, 2017. 155: p. 370–382.DOI: 10.1016/j.neuroimage.2017.04.046.

24. Kim, H., Suh, S., Joo, E. Y., and Hong, S. B., Morphological alterations in amygdalo-hippocampal substructures in narcolepsy patients with cataplexy. Brain Imaging Behav, 2016. 10(4): p. 984–994.DOI: 10.1007/s11682-015-9450-0.

25. Křečková, M., Kemlink, D., Šonka, K., et al., Anterior hippocampus volume loss in narcolepsy with cataplexy. J Sleep Res, 2019. 28(4): p. e12785.DOI: 10.1111/jsr.12785.

26. Juvodden, H. T., Alnæs, D., Lund, M. J., et al., Larger hypothalamic volume in narcolepsy type 1. Sleep, 2023. 46(11).DOI: 10.1093/sleep/zsad173.

27. Juvodden, H. T., Alnæs, D., Agartz, I., et al., Cortical thickness and sub-cortical volumes in post-H1N1 narcolepsy type 1: A brain-wide MRI case-control study. Sleep Medicine 2024.DOI: 10.1016/j.sleep.2024.02.031.

28. Iglesias, J. E., Insausti, R., Lerma-Usabiaga, G., et al., A probabilistic atlas of the human thalamic nuclei combining ex vivo MRI and histology. Neuroimage, 2018. 183: p. 314–326.DOI: 10.1016/j.neuroimage.2018.08.012.

29. Iglesias, J. E. Segmentation of thalamic nuclei. Laboratory for Computational Neuroimaging, Athinoula A. Martinos Center for Biomedical Imaging 2023 https://freesurfer.net/fswiki/ThalamicNuclei.[Accessed November 15 2024]

30. Iglesias, J. E., Van Leemput, K., Bhatt, P., et al., Bayesian segmentation of brainstem structures in MRI. Neuroimage, 2015. 113: p. 184–95.DOI: 10.1016/j.neuroimage.2015.02.065.

31. Olchanyi, M. Brainstem Substructures. 2024 https://surfer.nmr.mgh.harvard.edu/fswiki/BrainstemSubstructures.[Accessed November 15 2024]

32. Fischl, B., FreeSurfer. Neuroimage, 2012. 62(2): p. 774–81.DOI: 10.1016/j.neuroimage.2012.01.021.

33. American Academy of Sleep Medicine(AASM), International Classification of Sleep Disorders (ICSD) 3ed. 2014.

34. European Centre for Disease Prevention and Control, Narcolepsy in association with pandemic influenza vaccination - A multi-country European epidemiological investigation. 2012: European Centre for Disease Prevention and Control.

35. Hungnes, O., Iversen, B., Bergsaker, M., et al., Rapport om nytte av pandemivaksineringen, versjon 2. 2011, Norwegian Institute of Public Health

36. Juvodden, H. T., Alnaes, D., Lund, M. J., et al., Widespread white matter changes in post-H1N1 patients with narcolepsy type 1 and first-degree relatives. Sleep, 2018. 41(10).DOI: 10.1093/sleep/zsy145.

37. Juvodden, H. T., Alnaes, D., Lund, M. J., et al., Hypocretin-deficient narcolepsy patients have abnormal brain activation during humor processing. Sleep, 2019. 42(7).DOI: 10.1093/sleep/zsz082.

38. Anic-Labat, S., Guilleminault, C., Kraemer, H. C., Meehan, J., Arrigoni, J., and Mignot, E., Validation of a cataplexy questionnaire in 983 sleep-disorders patients. Sleep, 1999. 22(1): p. 77–87.

39. Heier, M. S., Evsiukova, T., Vilming, S., Gjerstad, M. D., Schrader, H., and Gautvik, K., CSF hypocretin-1 levels and clinical profiles in narcolepsy and idiopathic CNS hypersomnia in Norway. Sleep, 2007. 30(8): p. 969–73.

40. Knudsen, S., Jennum, P. J., Alving, J., Sheikh, S. P., and Gammeltoft, S., Validation of the ICSD-2 criteria for CSF hypocretin-1 measurements in the diagnosis of narcolepsy in the Danish population. Sleep, 2010. 33(2): p. 169–76.

41. Iglesias, J. E. Segmentation of hippocampal subfields and nuclei of the amygdala (cross-sectional and longitudinal). Laboratory for Computational Neuroimaging, Athinoula A. Martinos Center for Biomedical Imaging 2023 https://surfer.nmr.mgh.harvard.edu/fswiki/HippocampalSubfieldsAndNucleiOfAmygdala.[Accessed July 31 2024]

42. R Core Team. R: A Language and Environment for Statistical Computing. R Foundation for Statistical Computing. 2023 https://www.r-project.org/.[Accessed November 18 2024]

43. Winkler, A. M., Webster, M. A., Vidaurre, D., Nichols, T. E., and Smith, S. M., Multi-level block permutation. Neuroimage, 2015. 123: p. 253–68.DOI: 10.1016/j.neuroimage.2015.05.092.

44. Winkler, A. M., Ridgway, G. R., Webster, M. A., Smith, S. M., and Nichols, T. E., Permutation inference for the general linear model. Neuroimage, 2014. 92: p. 381–97.DOI: 10.1016/j.neuroimage.2014.01.060.

45. Benjamini, Y. and Hochberg, Y., Controlling the False Discovery Rate: A Practical and Powerful Approach to Multiple Testing. Journal of the Royal Statistical Society: Series B (Methodological), 1995. 57(1): p. 289–300.DOI: 10.1111/j.2517-6161.1995.tb02031.x.

46. Wada, M., Mimura, M., Noda, Y., et al., Neuroimaging correlates of narcolepsy with cataplexy: A systematic review. Neurosci Res, 2019. 142: p. 16–29.DOI: 10.1016/j.neures.2018.03.005.

47. Weng, H. H., Chen, C. F., Tsai, Y. H., et al., Gray matter atrophy in narcolepsy: An activation likelihood estimation meta-analysis. Neurosci Biobehav Rev, 2015. 59: p. 53–63.DOI: 10.1016/j.neubiorev.2015.03.009.

48. Dauvilliers, Y., Arnulf, I., Lecendreux, M., et al., Increased risk of narcolepsy in children and adults after pandemic H1N1 vaccination in France. Brain, 2013. 136(Pt 8): p. 2486–96.DOI: 10.1093/brain/awt187.

49. Partinen, M., Saarenpaa-Heikkila, O., Ilveskoski, I., et al., Increased incidence and clinical picture of childhood narcolepsy following the 2009 H1N1 pandemic vaccination campaign in Finland. PLoS One, 2012. 7(3): p. e33723.DOI: 10.1371/journal.pone.0033723.

50. Pizza, F., Peltola, H., Sarkanen, T., Moghadam, K. K., Plazzi, G., and Partinen, M., Childhood narcolepsy with cataplexy: comparison between post-H1N1 vaccination and sporadic cases. Sleep Med, 2014. 15(2): p. 262–5.DOI: 10.1016/j.sleep.2013.09.021.

51. Sarkanen, T., Alakuijala, A., Julkunen, I., and Partinen, M., Narcolepsy Associated with Pandemrix Vaccine. Curr Neurol Neurosci Rep, 2018. 18(7): p. 43.DOI: 10.1007/s11910-018-0851-5.

52. Sarkanen, T., Alakuijala, A., and Partinen, M., Clinical course of H1N1-vaccine-related narcolepsy. Sleep Med, 2016. 19: p. 17–22.DOI: 10.1016/j.sleep.2015.11.005.

53. Szakács, A., Darin, N., and Hallböök, T., Increased childhood incidence of narcolepsy in western Sweden after H1N1 influenza vaccination. Neurology, 2013. 80(14): p. 1315–21.DOI: 10.1212/WNL.0b013e31828ab26f.

54. Alakuijala, A., Sarkanen, T., and Partinen, M., Polysomnographic and actigraphic characteristics of patients with H1N1-vaccine-related and sporadic narcolepsy. Sleep Med, 2015. 16(1): p. 39–44.DOI: 10.1016/j.sleep.2014.07.024.

